# A multimodal investigation of perceptual awareness in Alzheimer’s disease

**DOI:** 10.64898/2026.08.27.26356661

**Authors:** Jonathan Huntley, Benjy Barnett, Daniel Bor, Marco Mancuso, Pedro A. M. Mediano, Lorina Naci, Stephen Fleming, Giacomo Bertazzoli, Linda Clare, Adrian M. Owen, Lorenzo Rocchi, Robert Howard

**Affiliations:** Department of Health and Community Sciences, University of Exeter, Exeter, UK; Division of Psychiatry, University College London, London, UK; Department of Experimental Psychology, University College London, UK; School of Psychological Sciences, Birkbeck, University of London, London, United Kingdom; Department of Psychology, University of Cambridge, Cambridge, UK; Department of Psychology, Queen Mary University of London; Centre for Brain and Behaviour, School of Biological and Behavioural Science, Queen Mary University of London; Human Neuroscience Department, Sapienza University of Rome, Rome, Italy; Department of Computing, Imperial College London, London, UK; School of Psychology, Trinity College Dublin, Global Brain Health Institute, Dublin, Ireland; Berenson-Allen Center for Noninvasive Brain Stimulation, Beth Israel Deaconess Medical Center, Boston, USA; Department of Physiology and Pharmacology, University of Western Ontario, London, ON, Canada; Department of Psychology, University of Western Ontario, London, ON, Canada; Institute of Neurology, University College London, London, United Kingdom; CIFAR Program in Brain, Mind and Consciousness, Toronto, ON, Canada; Institute of Cognitive Neuroscience, University College London, UK

**Keywords:** consciousness, awareness, Alzheimer’s disease, EEG, fMRI

## Abstract

Understanding how the contents of consciousness, particularly perceptual awareness, change with the progression of AD is crucial for meaningful person-centred care. This is especially important in severe AD when impairments in language and other cognitive domains limit reporting of experience.

We investigated whether electrophysiological (EEG) and fMRI signatures of perceptual awareness remain present in people with mild-moderate and severe AD. A visual masking paradigm examined visual awareness negativity (VAN) and late positive (LP) electrophysiological responses and activation in visual cortex and fronto-parietal regions; and a complex audio-visual (movie) task examined activation in fronto-parietal networks previously associated with perceptual awareness.

Healthy older controls demonstrated cortical responses characteristic of awareness in both EEG and fMRI modalities. In people with mild-moderate AD, VAN and LP markers and fronto-parietal activation were significantly reduced. In participants with severe AD, who were behaviourally minimally responsive, there was limited evidence of frontoparietal markers of perceptual awareness, however this may reflect attentional and task insensitivity.

These results demonstrate that the brain mechanisms associated with perceptual awareness become increasingly impaired with progression of AD. This motivates further investigation into the dimensions of awareness affected by the disorder with implications for treatment and management of people with dementia.

## Introduction

Dementia affects over 50 million people worldwide, with global prevalence growing exponentially^1^. Alzheimer’s disease, the commonest pathological subtype, accounts for 60-70% of dementia cases, and presents with progressive decline across multiple cognitive domains, impaired functioning and behavioural change^2^. Despite recent advances in amyloid-targeting treatments, there remains no means of preventing disease progression, so that a person living for 10 years with AD spends approximately 4 years in the advanced disease stage^3^.

Advanced dementia is characterised by severe cognitive impairment, including language impairment, and reliance on others for support of activities of daily living^4^. However, beyond established descriptions of loss of cognition and function, a central issue for people affected by AD, families and carers is the apparent loss of the self and potential loss of subjective phenomenological experience^5,6^. What makes AD such a feared disease is this loss of personhood; the loss of awareness of self and the environment, and the fear and uncertainty surrounding ‘what it is like’ to have advanced AD.

The question of ‘*what it is like’* to have dementia is commonly asked by patients and caregivers and reflects a primary question in consciousness science. Influential frameworks for consciousness science have described and differentiated the *level* of consciousness (on a scale from unconsciousness to wakefulness), and the *contents* of consciousness, (what we are aware of)^7^. There is evidence that changes in the contents of awareness occur early in AD and progress with worsening disease severity. Impaired awareness of cognitive and functional deficits is characteristic of mild-moderate dementia caused by AD and is associated with neurodegeneration in brain regions and networks that support metacognition and self-awareness^6,8,9^. However, there remains uncertainty about which specific elements of awareness decline in AD. Clinically, people with advanced AD appear unaware of themselves and their surroundings and often do not recognise familiar loved ones^10^. Loss of facial recognition (prosopagnosia) is characteristic of AD^11^. However, a more fundamental issue than impaired visual memory or recognition is whether a person with advanced AD may have more generally impaired awareness of sensory information from their environment. Although there are consistent patterns to neurodegeneration in AD progression, there is also significant variability in clinical presentation and course. Therefore, it is likely that there will be a variability in capacity for, and content of, awareness between individuals with advanced AD. This highlights the importance of understanding how the content of consciousness may change with AD in efforts to provide meaningful person-centred care to patients.

Studying the content of consciousness and its neural correlates in healthy adults usually involves the collection of subjective reports of experience, for example using psychophysical paradigms that contrast visible and invisible stimuli and asking participants whether they have seen a stimulus or not. This often uses visual masking paradigms, in which the target picture (e.g. a face) is displayed very briefly, immediately followed by a second picture (a mask) which removes the target stimuli from awareness^12^. Previous research has combined visual masking paradigms with electroencephalography (EEG) and functional magnetic resonance imaging (fMRI) to identify signatures of conscious perception^13^. In event-related potentials (ERP) studies, visual awareness negativity (VAN) and late positivity (LP) are important markers of conscious visual perception^14^. The VAN is a negative deflection seen over posterior electrode sites that typically occurs at around 200-300 ms and is considered an early marker of conscious perception^15–17^. The LP is a positive deflection seen in posterior electrodes 300-500 ms after the stimulus and is considered to reflect the availability of information to cognitive systems to control reasoning and behaviour^15–17^. In fMRI studies, activation in the visual cortex and fronto-parietal regions has been associated with visual awareness^13,18,19^. The neural correlates and theoretical underpinnings of consciousness remain an area of active research^20^. However, the reliability of these validated paradigms to produce ERP or fMRI signatures that covary with visual awareness in people who can report their conscious experience have led to extrapolation to assessment of perceptual awareness in clinical groups who are unable to provide subjective verbal reports. These “no-report” paradigms have been used to investigate neural correlates of consciousness in preverbal infants^21^, in adults during anaesthesia and sleep^22^, and in patients with disorders of consciousnes^19^. Using these no-report paradigms therefore provides an opportunity to investigate changes in conscious perception in people with AD who are unable to reliably report their experience. In this study we operationalise perceptual awareness as the presence of these ERP and fMRI signatures of visual awareness validated in previous clinical groups. Using validated visual masking paradigm and movie tasks, our aim was to assess visual awareness in people with AD, and answer the following research questions:

1) In response to visual stimuli, are ERP signatures of visual awareness (VAN/LP) reduced in AD compared to healthy older control participants?
2) Is there fMRI evidence of brain activity that goes beyond primary visual areas and activates a fronto-parietal network, reflecting preservation of higher-level awareness in AD?
3) Is there evidence for reduction in markers of visual awareness with progression of AD into the advanced stages?

## Materials and methods

### Participants

Participants were recruited from NHS memory services (North London NHS Foundation Trust) and the UK national Join Dementia Research database. Patients with AD had diagnoses made prior to the study by clinical services according to ICD-10 criteria^23^. AD severity was classified using the clinical dementia rating scale (CDR-SOB)^24^ and Global deterioration scale (GDS)^4^. Older adult participants without dementia (healthy older controls) were also recruited. Exclusion criteria included any concurrent medical condition, psychiatric illness or medication that may interfere with awareness, contraindications to fMRI or significant visual or hearing impairment. All participants were assessed for capacity to provide informed consent. As participants with severe AD lacked capacity to consent, following the legal framework of the Mental Capacity Act (2005), personal consultees were identified and asked to provide a declaration that the person would have wished premorbidly to take part in the study^25^. Informed consent was therefore obtained from all participants and/or their legal guardian(s). The study was performed in accordance with the Declaration of Helsinki and approved by the Wales 6 NHS Research Ethics Committee (18/WA/0012).

### Assessing conscious visual perception

In experiment 1, a visual masking paradigm was used with EEG and *a priori* event related potential (ERP) signatures of visual awareness were assessed, specifically the occipital VAN and LP^15,26^. In experiment 2, the same visual masking task was used during fMRI. In experiment 3, a naturalistic movie watching task was used during fMRI^19,26^. In the fMRI experiments *a priori* regions of interest included occipital cortex, and fronto-parietal regions previously validated as markers of awareness using the same task in healthy populations and patients with disorders of consciousness^19,27^.

### Visual-masking paradigm

A visual masking paradigm was adapted from a study of visual awareness in infants, who, like people with severe AD, are unable to report their conscious experience^21^. The task consists of a series of pictures, either of neutral faces (stimuli) or scrambled faces (masks), presented for a duration of either 33 ms or 200 ms. Previous studies have demonstrated that stimuli presented for 200 ms are consciously perceived, whereas stimuli for 33 ms are subliminal^28^. Trials began with a fixation cross for 1000 ms. The critical stimuli either consisted of a face or a mask and were then presented for either 33ms (subliminal condition) or 200 ms (visible condition). The critical stimuli were flanked by a forward mask (300 ms) and a backward mask (33 ms), followed by a final mask (1500 ms). During the ERP experiments, 10 pseudo-random blocks of 20 trials (5 trials of 4 trial types) were presented. During the fMRI experiment, 180 trials were presented in a random order across 3 scanning runs, with 45 trials per condition (visible-face, subliminal-face, visible-mask, subliminal-mask). Participants were instructed to passively attend to the stimuli, and infra-red eye tracking was used to ensure their eyes were open and they were watching the screen throughout the experiment.

### Movie task

A naturalistic movie watching task was used based on previous studies^19,27^. An edited 8-min sequence of the TV series episode, “Alfred Hitchcock Presents - Bang! You’re Dead” was presented. Participants were asked to simply watch the film, and an infrared camera placed inside the scanner was used to ensure participants maintained eye opening and focused gaze on the screen during the movie. Noise cancellation headphones were used for sound delivery. Previous studies using this movie task have demonstrated synchronised activity in fronto-parietal regions in healthy controls that is associated with conscious awareness of the movie. The within- and between-participant reproducibility of this fronto-parietal activity allows it to be used as a no-report marker of awareness of the movie in clinical groups^19,27^. If a patient’s activity in frontal and parietal regions is tightly correlated with that of healthy participants over time, such functional correspondence can be interpreted as demonstrating alignment in awareness during movie-viewing at a single-subject level^19^.

### ERP acquisition and analyses

EEG signal was recorded from 63 active electrodes (actiCAP), positioned according to the international 10-10 system, referenced online to Oz with ground at Fpz. Impedances were kept below 5 kΩ and sampling frequency was 5000 Hz. Preprocessing was performed with EEGLAB (v14.1.1, MATLAB vR 2018b). Data were epoched (−1.7 to 1.7s) using a baseline from −1200 to −400ms, to avoid contamination from responses to the fixation mask. Signals were band-pass (0.1-100 Hz), and band-stop (48-52 Hz) filtered using a 4^th^-order Butterworth filter, downsampled to 1000Hz, then further epoched (−1 to 1s) to minimise edge artefacts. Noisy epochs were rejected following visual inspection. Independent component analysis (fastICA) was used to remove ocular and muscle artefacts and data were re-referenced to the common average. In line with Kouider et al. (2013), analyses were focused on an occipito-parietal cluster (O2-Oz-O1-POz-PO3-PO4-PO8-PO7)^21^. Mean voltage across these electrodes was computed, and peak amplitudes corresponding to the minimum for VAN and maximum for LP within each corresponding time range (250-350 and 350-550 ms, respectively) were extracted. A frontal component, N400, corresponding to the minimum peak amplitude between 350-550 ms, was also assessed as an index of cognitive processes including attention, memory and comprehension^29–32^. These were then compared between visible and subliminal face trials using a mixed model ANOVA to examine main effects of group, component and group x component interactions. Post-hoc comparisons between groups were assessed as appropriate using t-tests adjusted for multiple comparisons using the Holm-Bonferroni procedure^33^.

### fMRI acquisition and analyses

Scanning was performed on a 3T Siemens Prisma with a 20-channel head coil. Structural images were acquired using an MPRAGE sequence (1×1×1mm voxels, 176 slices). Field maps were obtained with a double-echo FLASH (gradient echo) sequence with TE1 = 10ms and TE2 = 12.46ms (64 slices, slice thickness = 2 mm, gap = 1 mm, in plane FoV = 192 x 192 mm, resolution = 3 × 3 mm). For the visual masking paradigm, functional images were acquired using a 2D EPI sequence (3 mm isotropic voxels, TR = 3.36s, TE = 30 ms, 48 slices). For the naturalistic movie paradigm, functional images were acquired in the same session using a 2D EPI sequence (3 mm isotropic voxels; TR = 2.10s; TE = 30 ms, 30 slices). Preprocessing was performed with SPM12 (Statistical Parametric Mapping; www.fil.ion.ucl.ac.uk/spm) using standard procedures^34–36^: removal of first 5 volumes, realignment and unwarping using local field maps^37^, slice-timing correction^38^, segmentation of structural images^39^, normalization to MNI space and spatial smoothing (8mm FWHM). Motion thresholds were 3 mm translation and 1 degree rotation. Preprocessing and construction of first- and second-level models used standardized pipelines and scripts available at https://github.com/metacoglab/MetaLabCore/.

### General Linear Model (GLM)

For the visual masking paradigm, general linear models included four regressors of interest per block (visible-face, subliminal-face, visible-mask, and subliminal-mask), modelled as stick functions at stimulus onset and convolved with the canonical haemodynamic response function. Motion parameters were included as nuisance regressors, and low-frequency drifts were removed with a standard 1/128Hz high-pass filter. For control and AD groups, whole-brain single-subject contrast images assessing face sensitivity (faces > masks) and visual awareness (visible-faces > subliminal-faces) were entered into second-level random-effects analyses. One-sample *t* tests were used to assess within-group statistical significance and two-sample independent *t* tests were used to test for differences between the control and AD groups. For severe AD patients, owing to the small number of patients, only single-subject contrast images were computed. To reduce the risk of false negatives when assessing for markers of awareness in people with AD, univariate contrasts were initially examined at a liberal voxelwise threshold of p < .05 uncorrected, with an arbitrary cluster forming threshold of 250 for the purposes of visualisation. As such, results from univariate contrasts were conducted to provide only qualitative illustrations of neural responses to conscious and unconscious stimuli, while multivariate decoding analyses (below) provide valid whole-brain statistical inference.

### Multivariate Decoding

To demonstrate statistical differences in activation associated with visual awareness, we performed whole-brain searchlight decoding analyses. Decoding analyses are sensitive tools used to identify multivariate neural patterns associated with specific cognitive processes and have been widely used to characterise multivariate neural patterns associated with awareness^40–42^. Beta coefficients were computed per trial^43^ and used to train a binary LDA decoder to distinguish visible from subliminal-face trials. Searchlights (radius = 4 voxels, total = 257 voxels) moved throughout the brain, with each voxel included as the centre voxel in a searchlight. The decoder was trained with a 5-fold cross-validation with balanced trial numbers. The accuracy of each searchlight’s decoder was averaged across folds, and this value was stored at the centre of the searchlight, producing a whole brain map of decoding accuracy. Above chance accuracy of such a decoder indicates that neural activity associated with a participant’s level of awareness exists within the searchlight, and as such this procedure can be used to examine which neural regions are associated with visual awareness. All decoding analyses were performed using custom MATLAB (2021b) scripts. Significance was assessed using permutation-based inference^44^. Class labels were permuted 25 times per participant and bootstrapped to generate group-level null distributions (10,000 samples). Observed maps were compared with null distributions, with results thresholded at *p* < .05 (FDR corrected using the Benjamini–Hochberg procedure).

### Movie paradigm time series analysis

Synchrony analyses of the fMRI time-series assessed awareness-related engagement with the movie by correlating each participant’s BOLD time course with the mean time-course of other participants. Similar to previous studies using this paradigm, fronto-parietal synchrony is taken as a marker of awareness and engagement with the movie^19^. In the case of between-group comparisons between patients with mild-moderate AD and healthy controls, the time-course of each patient was correlated with the mean time-course of all healthy controls combined. This procedure resulted in maps of correlation coefficients for each participant, which were tested against zero at the group-level using one-tailed tests and cluster-level FWE correction (*p* < .05). Single subject analyses of patients with severe AD used the same procedure, with the time-course of each patient correlated with the mean time-course of all healthy controls combined, resulting in single subject maps of correlation coefficients which were converted to *t* values (df = number of timepoints – 2) and FDR-corrected for multiple comparisons (*q* = 0.01, *p* < .05).

## Results

### Event Related Potentials

The ERP study involved 28 controls, 17 participants with mild-moderate AD and 7 with severe AD (Table 1). The main contrast compared ERPs to faces presented long enough for conscious perception (200 ms) versus subliminal presentations (33 ms). Two established ERP signatures of conscious perception were examined: visual awareness negativity VAN and the LP components across occipital regions. A frontal component (N400) was also assessed^29,30^.

**Table 1:** Demographic and cognitive data on participants.

|  | Control (n= 28) | Mild-moderate AD (n = 17) | Severe AD (n=7) |
| --- | --- | --- | --- |
| AGE (SD) | 75.43 (5.63) | 76.59 (6.65) | 83.14 (6.20) |
| MALE/FEMALE | 10/18 | 8/9 | 2/5 |
| CDR (median (IQR)) | 0 (0) | 4.5 (6) | 18 (1) |
| GDS (median (IQR)) | 1 (0) | 4 (2) | 7 (1) |
| sMMSE | 29.60 (0.60)* | 22.53 (6.17) | 2.83 (4.92) |

*CDR = Clinical Dementia Rating Scale; GDS = Global Deterioration Rating Scale; sMMSE = standardised Mini Mental State Examination; \*n = 20 as sMMSE conducted with controls later in the course of the study.*

### ERP signatures of visual perception are reduced in AD

Healthy controls showed robust VAN and LP components (Figure 1). ANOVA revealed significant group differences for both VAN (F(2,49) = 5.46, p = 0.007) and LP (F(2,49) = 13.42, p = 0.00002). Compared to controls, the mild-moderate AD group showed significantly reduced amplitudes of both components (VAN: T = −2.46, p = 0.03; LP: T = 4.13, p = 0.0003). Significant differences were also observed between AD groups, (VAN: T = −2.65, p = 0.03; LP: T = 3.19 p = 0.005) with only negligible VAN and LP responses in severe patients. The amplitude of both VAN and LP components correlated with disease severity (VAN: *r*=0.441, p=0.001; LP: *r*=-0.565, p<0.001; Figure 2). Analyses of the frontal N400 component also showed group differences (Figure 2, B and C); however, this component is not exclusively implicated in visual awareness but is commonly reduced in dementia and associated with processing of stimuli in multiple modalities and across multiple cognitive domains^29–31^.

**Figure 1:**
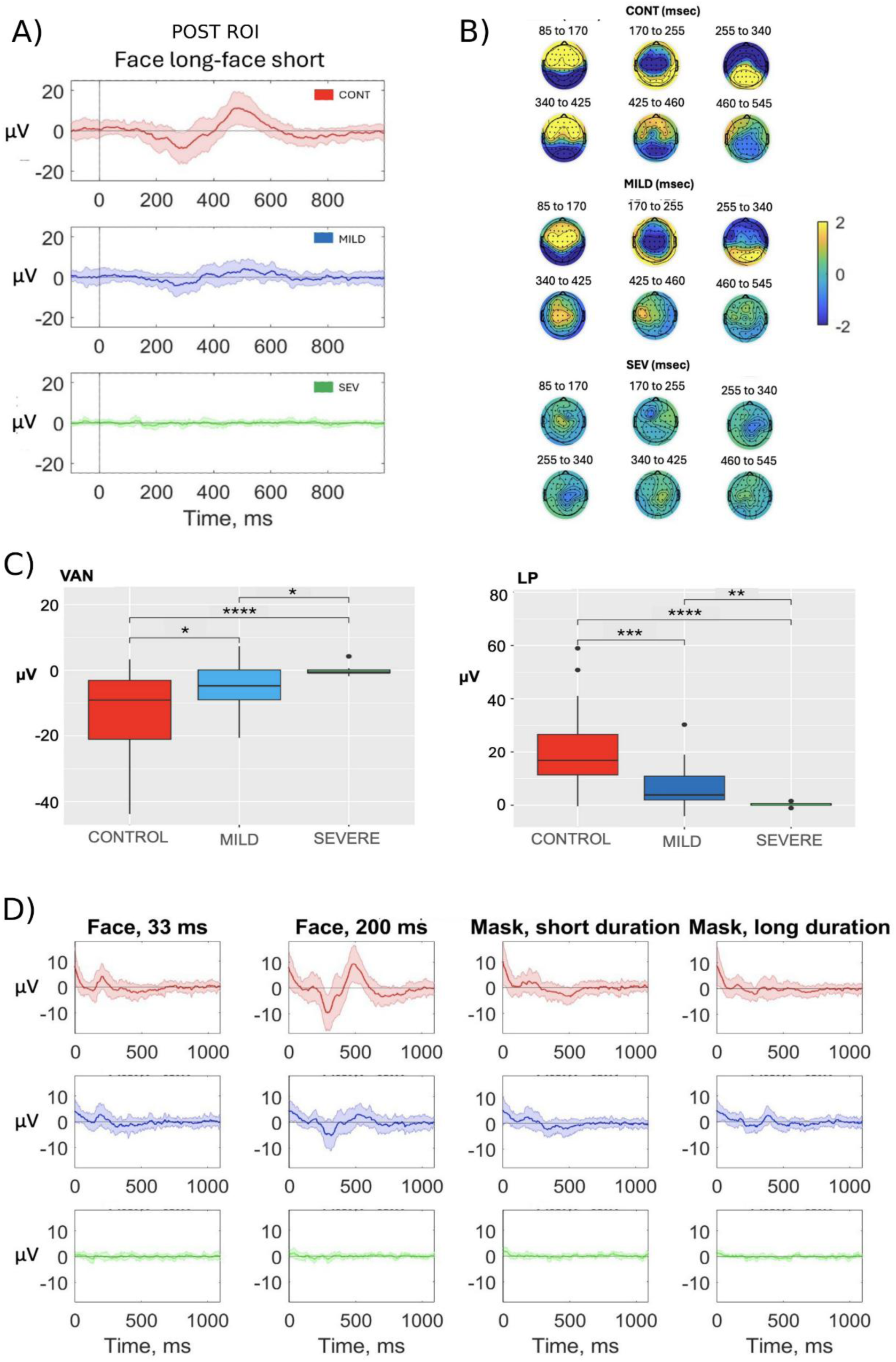
ERP markers of visual perception. A) Amplitude-time plots of visible – subliminal face contrasts averaged across 8 occipital parietal regions, for control group (red), mild-moderate AD group (blue) and severe AD group (green) (means and SEM). B) Topographic maps of the ERP response to the visible-subliminal face contrast in the control (top), mild-moderate AD (middle) and severe AD (bottom) groups. C) Statistical comparison of VAN and LP components for each group reveal significant group differences * p < 0.05, ** p < 0.01, *** p<0.001,**** p<0.0001 (central line = median, box = IQR, whiskers extend to data points within 1.5 × IQR from the quartiles) D) Amplitude-time plots for all conditions, for control group (red), mild-moderate AD group (blue) and severe AD group (green) (means and SEM).

**Figure 2:**
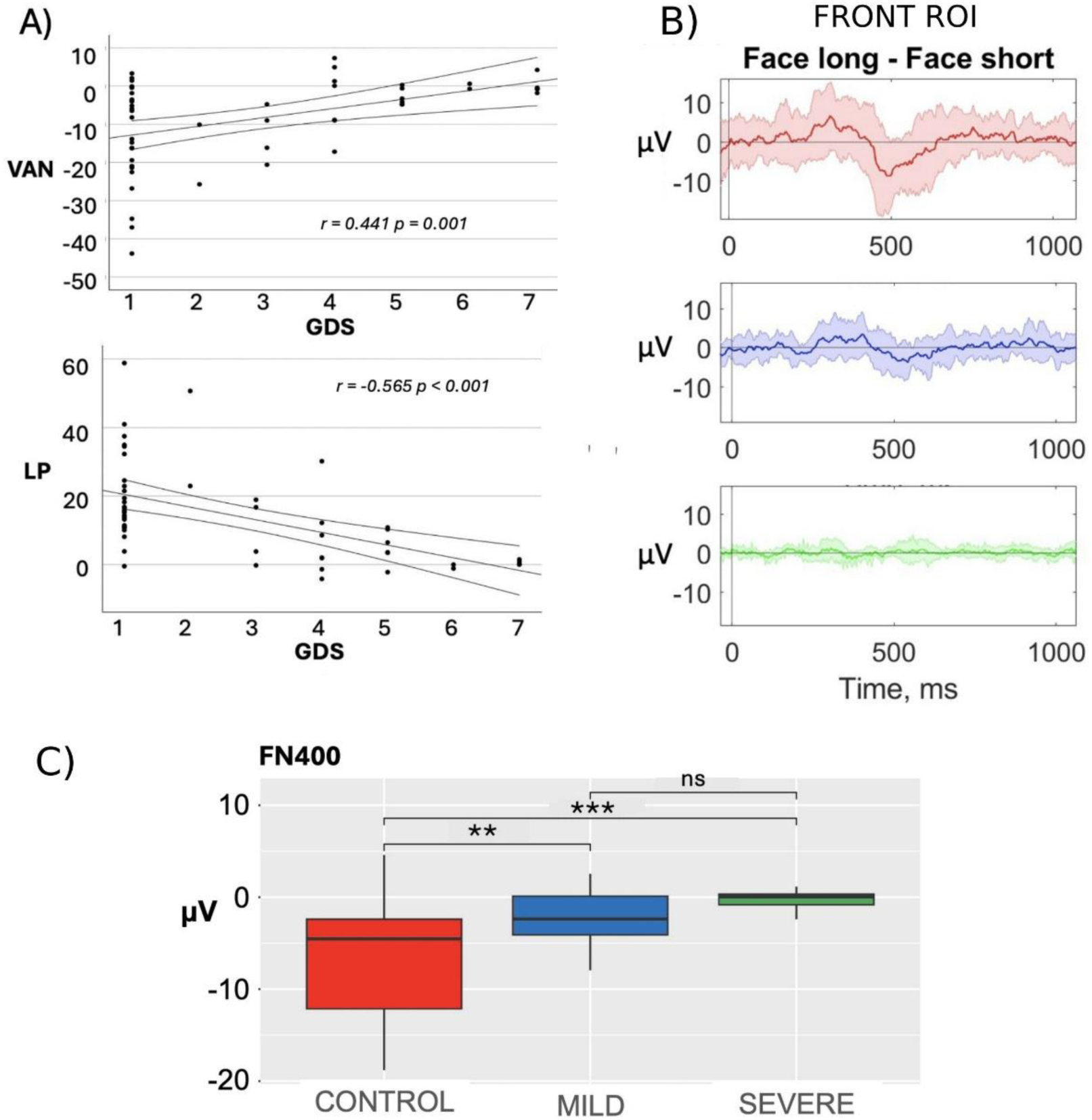
ERP frontal markers and correlation with disease severity. A): correlation of VAN and LP with dementia severity (GDS= Global deterioration rating scale); (solid bold line = fitted linear regression line, surrounding curved lines = 95% CI for the regression estimate). B) Amplitude-time plot averaged across frontal electrodes for control group (red), mild-moderate AD group (blue) and severe AD group (green). C) statistical comparison of frontal N400 component for visible-subliminal face contrast, revealing significant differences between AD groups and healthy controls. ** p < 0.01, *** p<0.001, ns = non-significant

Although ERP signatures of visual perception remained detectable, albeit reduced, in mild-moderate AD, including components associated with unconscious processing (e.g. P100: Figure 1D), severe AD participants exhibited minimal response across all conditions. The lack of components associated with both unconscious and conscious processing may suggest a general lack of sensitivity to the task, rather than a selective loss of awareness-related processing. This prompted further investigation of task sensitivity and awareness-related processing during the fMRI experiment

### fMRI

The fMRI study included 26 controls, 14 patients with mild-moderate AD and 4 with severe AD. The reduced number of participants reflected contraindications and reduced tolerability to fMRI particularly in the AD participants. Due to the reduced number of severe AD participants, we report the group comparisons between controls and mild-moderate AD participants and present the severe AD data as a case series.

### Faces evoke widespread activation in controls and patients

To ensure participants were sensitive to the target face stimuli presented in the experiment, we trained a decoder to decode visible face stimuli from masks. In healthy controls, decoding was successful across visual, parietal and frontal cortices (Figure 3, top; Supplementary Table 1). AD patients also showed successful decoding throughout visual, parietal, and frontal regions (Figure 3, middle; Supplementary Table 1), suggesting that during passive viewing, AD patients were sensitive to the experimental paradigm and face stimuli. Controls showed greater sensitivity to faces in small regions of the visual and parietal cortex (Figure 3, bottom; Supplementary Table 2), but no difference was found in frontal regions.

**Figure 3:**
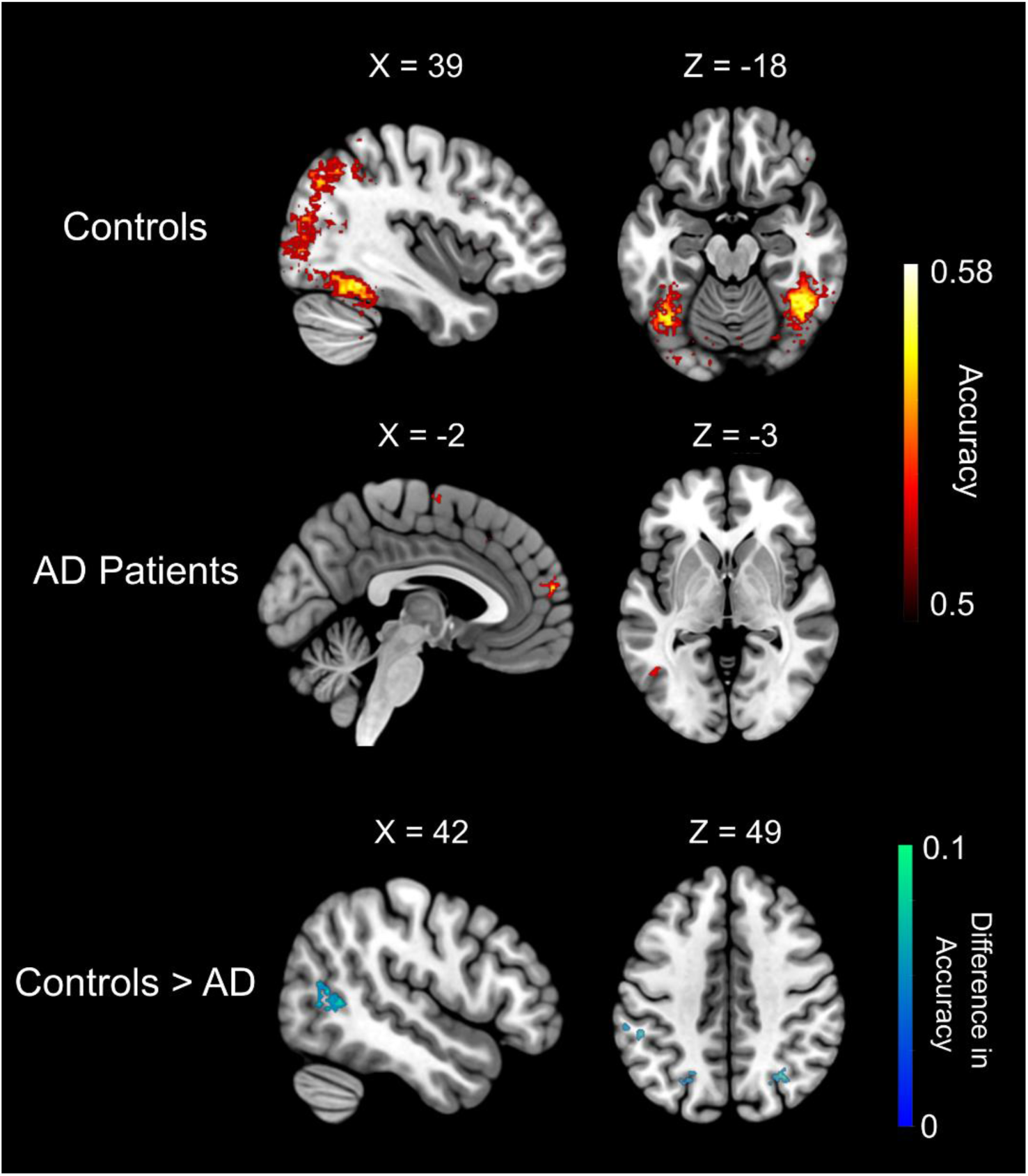
Healthy controls and AD patients both show sensitivity to visible faces. Searchlight decoding analyses reveal regions across occipital, fusiform and medial pre-frontal regions where face information could be decoded, indicating that face stimuli were processed by participants. Healthy controls showed significantly better face decoding in regions of the visual and parietal cortex, but no difference was found in frontal cortices. Maps are thresholded at p<.05, corrected for multiple comparisons. Clusters are presented in Supplementary Table 1 and 2.

### Univariate analyses of awareness

Whole brain univariate analyses comparing visible versus subliminal faces were conducted at a liberal p< 05 uncorrected threshold to provide a qualitative visualisation (Supplementary Figure 1, Supplementary Table 3). Univariate analyses revealed increased fusiform gyrus and dorsolateral prefrontal cortex (dlPFC) activity in controls, (Supplementary Figure 1; top; Supplementary Table 3), with reduced activation across motor and prefrontal regions in the AD group (Supplementary Figure 1; middle). Direct comparison between the control and AD groups showed greater awareness-related activation in controls versus AD patients in the left fusiform gyrus, right anterior insula and right dlPFC (Supplementary **Error! Reference source not found.**; bottom; Supplementary Table 3), however these contrasts provide only qualitative illustrations while multivariate decoding analyses (below) provide valid whole-brain statistical inference.

### Decoding analyses reveal diminished awareness-related activity in AD

To provide a more sensitive multivariate analysis of awareness-related activity, searchlight decoding analyses were performed throughout the entire brain to identify multivariate neural patterns that correlated with visual awareness. Searchlight decoding showed significantly above chance classification of visible versus subliminal trials across visual, parietal, and frontal regions in controls (Figure 4, top; Supplementary Table 4). In AD patients, decoders were also able to detect awareness-related activity across fusiform and prefrontal regions (Figure 4, middle; Supplemental Table 4), although decoding accuracy was significantly better for healthy controls throughout visual, parietal, and prefrontal regions (Figure 4, bottom; Supplementary Figure 3). These decoding results are in keeping with our univariate contrasts indicating a reduction in neural responses associated with visual awareness in AD in both occipital and fronto-parietal regions, with fronto-parietal activity more significantly reduced in AD.

**Figure 4.**
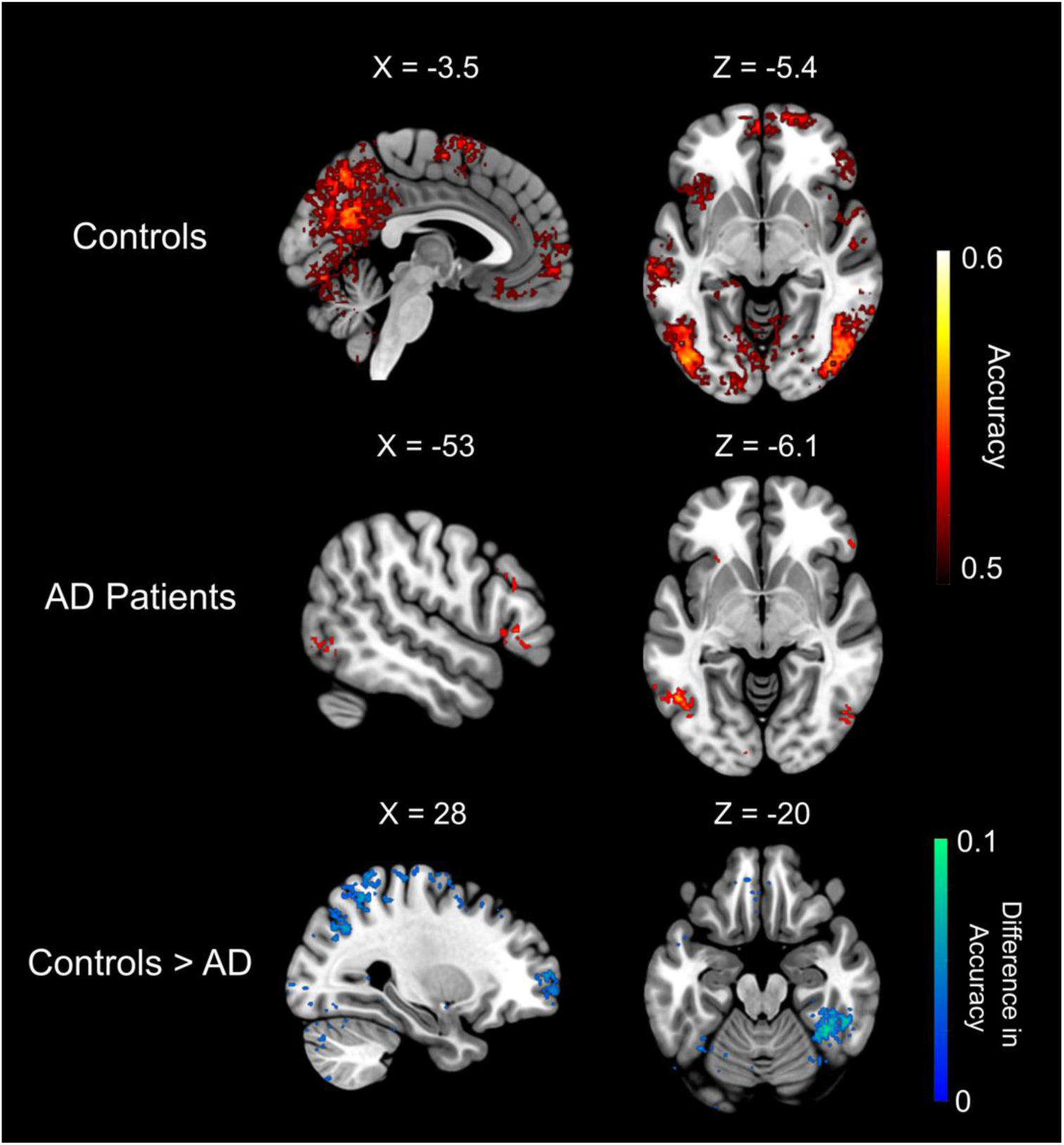
Decoding analyses reveal existing, diminished, awareness-related activity in AD. Searchlight decoding analyses were conducted with decoders trained to classify visible and subliminal face trials in searchlights throughout the brain. In healthy controls, the decoder could classify visible from subliminal trials significantly above chance throughout visual, parietal, and frontal regions (top). In AD patients, decoders were also successful in identifying neural patterns associated with awareness in visual and prefrontal regions (middle). In keeping with univariate analyses, there was a significant increase in decoding accuracy for controls vs. AD patients, indicative of diminished neural markers of awareness in AD. Maps are thresholded at p < 0.05 corrected for multiple comparisons. Clusters are reported in Supplementary Table 4 and Supplementary Table 5.

### Neural responses to visible stimuli in advanced AD

Single-subject analyses in severe AD revealed no significant awareness-related activity in all but one patient, even at the lenient statistical threshold of *p* <.05, uncorrected (Figure 5; *Patients 2-4*). One participant, however, exhibited a classic ‘ignition-like’ neural profile associated with a contrast of visible and subliminal faces, extending across bilateral fusiform, parietal, and medial and lateral pre-frontal cortices (Figure 5 *patient 1*; Supplementary Table 6). Of note, observations of behaviour showed that this participant was less impaired than the others in the severe group (GDS 6 vs GDS 7) and remained capable of some verbal communication.

**Figure 5.**
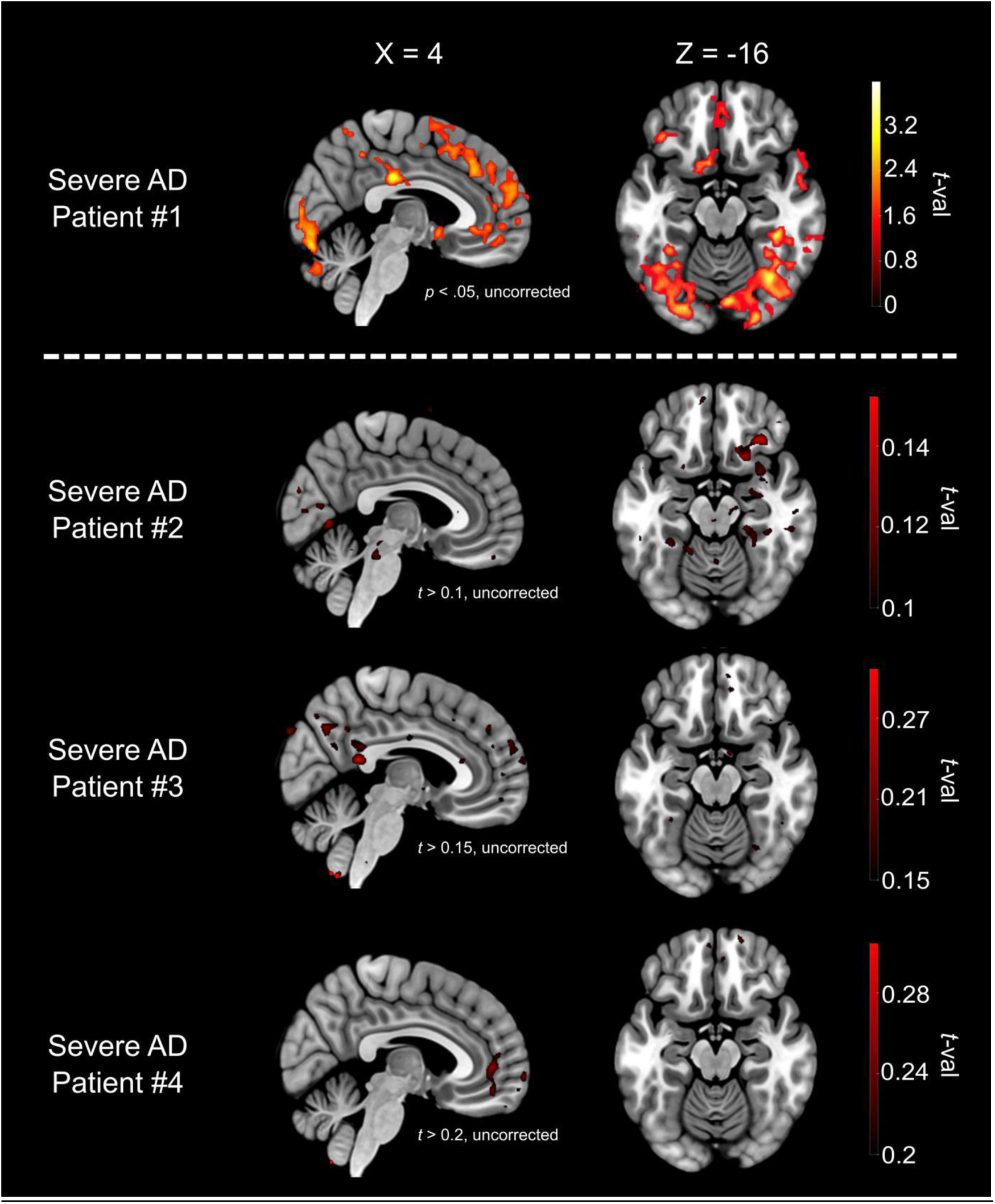
Participants with severe AD. For single subjects with severe AD, contrasting trials where patients viewed visible faces vs. subliminal faces illustrates the loss of neural markers of awareness in most patients (#2 – #4). For visualisation purposes, maps for these patients are presented at a threshold of t > 0.1. One patient with severe AD (#1) showed significant awareness-related activity across posterior and frontal regions. Notably, this patient was the least severe (GDS 6) and had some residual communication ability. The map for patient #1 is presented at p <.05, corrected for multiple comparisons.

### Null results in severe patients may be due to general insensitivity to the stimuli

To assess whether an absence of clusters in these single-subject qualitative maps was unique to participants with severe AD, we extracted functional regions of interests (ROIs) that exhibited sensitivity to the visibility of face stimuli in healthy controls. Two ROIs were created, first, an ROI that comprised of the right fusiform cluster, second, a functional ROI which spanned lateral and medial portions of the PFC. All but one severe AD patient registered t-values near zero in the right fusiform (Supplementary Figure 2, top-left) and PFC (Supplementary Figure 2, top-right), suggesting no increase in activity for visible versus subliminal faces in these patients. However, across both ROIs, the Control and Mild AD groups also contained single subjects who failed to exhibit an increase in activity for visible versus subliminal faces. As such, despite exclusively finding ignition-like activity in the only severe AD patient to exhibit residual communicative abilities, the null findings in the other severe AD patients cannot conclusively be attributed to an absence of such abilities, as null results also occur in a proportion of healthy individuals (Supplementary Figure 2, top). Additionally, within the same ROIs, the t-values computed from a visible face versus scrambled mask contrast were clustered around 0 for all three patients with severe AD who showed no response to the visibility of faces (Supplementary Figure 2; bottom), suggesting a global failure to respond to the faces stimuli during the masking task rather than a specific dissolution of awareness-related neural activity.

### Frontoparietal activity is also reduced in AD during complex audiovisual stimuli

Markers of awareness to more complex audio-visual stimuli were assessed using the movie watching task. For one patient with severe AD, excessive movement towards the end of the movie meant it was only possible to analyse the first 200 TRs of their data. However, this still amounted to over 80% of the movie duration.

Examination of within-subject correlations in the control group demonstrated widespread sensory and frontoparietal synchrony associated with conscious engagement with the movie (6A). In contrast, mild-moderate AD participants showed within-group synchrony, largely restricted to posterior regions (Figure 6B). The mild-moderate AD group displayed synchrony in auditory and visual regions with the control group, but limited fronto-parietal synchrony (Figure 6C). In contrast, of the severe patients, three participants elicited no significant synchrony with the control group in any regions, and one participant, who had also displayed awareness-related activation in the faces task, demonstrated visual and auditory synchrony with the control group, but no fronto-parietal synchrony (Figure 6D). These findings indicate disruption in AD of frontoparietal networks that have been associated with awareness of and engagement with the movie stimuli in healthy adults, particularly in the advanced stages of AD.

**Figure 6.**
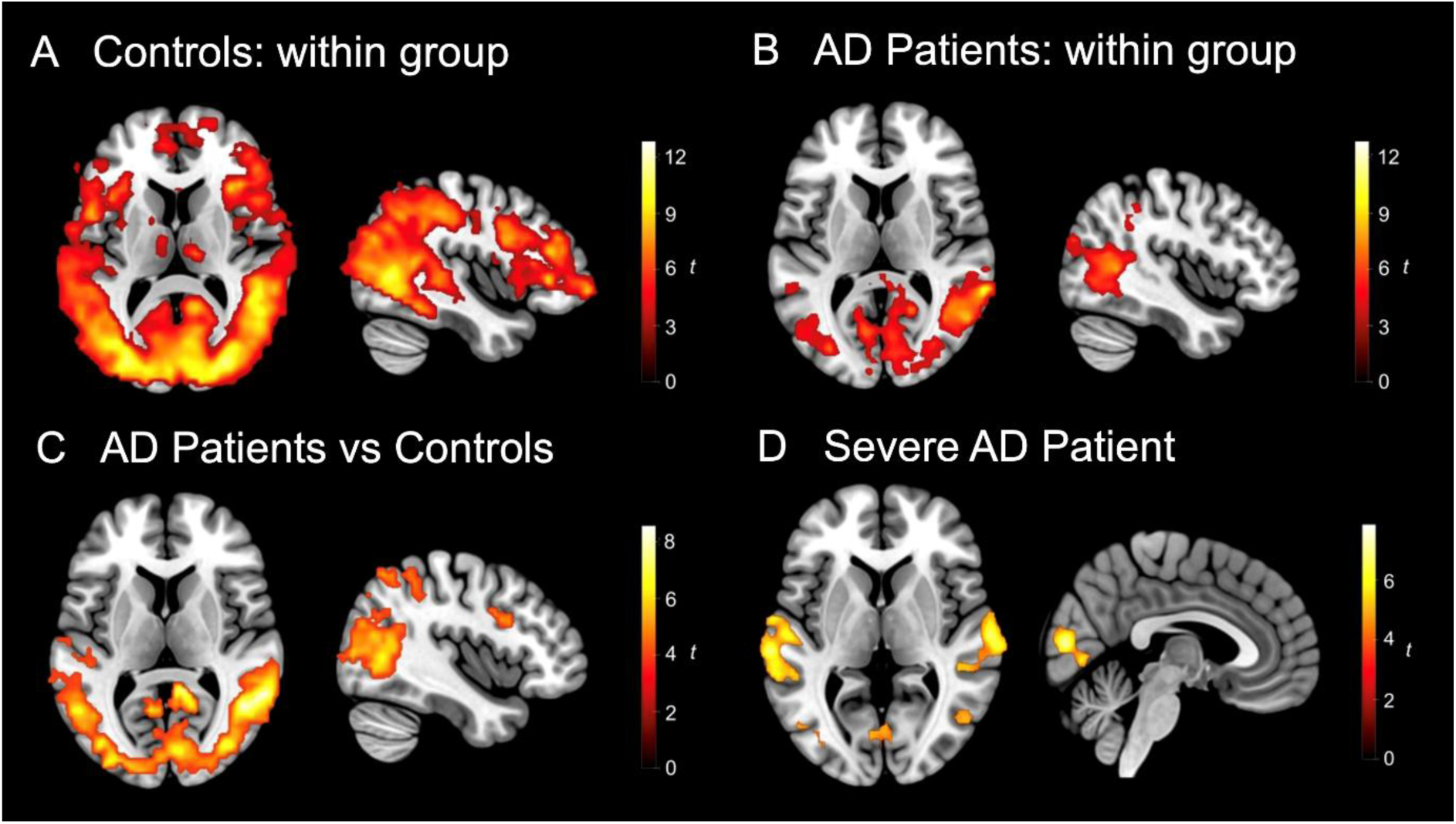
Synchrony during movie task. A) Control participants demonstrate significant within group synchrony in frontoparietal, auditory and visual regions; B) Mild-moderate AD participants demonstrate significant within group synchrony predominantly within auditory and visual regions; C) Comparison of synchrony between mild-moderate AD and controls demonstrates synchrony in auditory and visual regions with the control group, with limited fronto-parietal synchrony; D) correlation map of the single severe AD patient who showed any synchrony with the healthy controls, demonstrating only synchrony in sensory regions, with no frontoparietal synchrony. The three other severe AD patients showed no synchrony with healthy controls. Please see supplementary tables 7-10 for further information of regions showing significant correlations.

## Discussion

This study provides converging EEG and fMRI evidence that neural markers of perceptual awareness are progressively reduced across the range of severity of AD. Using complementary paradigms and multiple candidate markers of visual awareness, we show that while individuals with mild–moderate AD retain sensitivity to faces, neural signatures associated with conscious visual perception are significantly reduced relative to healthy controls. In the visual masking paradigm, reduced awareness in mild–moderate AD was reflected by attenuated VAN and LP components and reduced activation and decoding of stimulus visibility across occipital and fronto-parietal regions. Similar reductions in synchronised fronto-parietal activity were observed during naturalistic movie viewing, suggesting diminished conscious engagement with dynamic visual input. Together, these findings suggest that in mild-moderate AD there is impairment in awareness-related neural processes. . In patients with severe AD, awareness-related markers were largely absent, with residual frontal activity observed only in one individual who retained some communication abilities. However, null results in the majority of severe AD participants may reflect attentional and task insensitivity rather than selective impairment in awareness-related processes.

Our results are consistent with theoretical accounts of consciousness including the Global Neuronal Workspace Theory (GNWT), and Higher-Order theories (HOT). GNWT proposes that conscious perception depends on the propagation of sensory information from posterior cortices to fronto-parietal networks^13,22^. HOT propose that fronto-parietal networks are required for monitoring of sensory information^45^. Neurodegeneration in AD disproportionately affects large-scale cortical networks implicated in awareness, self-monitoring, and metacognition, including fronto-parietal and default mode networks^9,46,47^. Disconnection within these systems as AD progresses may reduce the likelihood that sensory representations achieve the stability or precision required for global ‘ignition’, leading to weakened or inconsistent frontal responses to visible stimuli. Our finding of reduced decoding of perceptual content in visual cortex further suggests that sensory representations themselves may also be degraded, limiting their capacity to enter conscious access.

Although univariate fMRI responses in early visual cortex appeared similar between groups, multivariate fMRI and ERP analyses revealed reduced posterior markers of stimulus visibility in AD, possibly indicating reduced representational precision. This highlights the importance of multivariate and temporally sensitive measures when assessing awareness-related processes^40,42^. Future studies incorporating direct measures of functional connectivity during near-threshold perception may help disentangle degraded sensory encoding from impaired global broadcasting. Longitudinal studies would also help elucidate changes in functional connectivity and posterior and fronto-parietal markers of awareness within individuals as AD progresses.

A key limitation in the study is the absence of trial-by-trial subjective reports from participants, particularly in those with advanced AD. However, the paradigms and neural markers employed were specifically chosen as ‘no-report’ paradigms and have been robustly linked to subjective visual awareness across several decades of research, including studies of infants and disorders of consciousness^18,19,21^. Using no-report paradigms allows comparison across groups, including non-communicative patients with severe AD. While an inference gap remains in non-communicative patients, the strong coupling between these markers and reported experience in other populations provides a principled basis for cautious inference. Importantly, this study does not attempt to identify necessary or sufficient neural correlates of consciousness but rather examines whether established markers associated with perceptual awareness are systematically reduced in AD and with disease progression.

Attentional and cognitive impairments represent a significant potential confound, with the association between awareness, attention and other cognitive domains remaining an area of research and debate^48–51^. However, some theories suggest that attention is necessary for consciousness^52^ and that attention and consciousness are inextricably linked^53^. In healthy controls and mild–moderate AD patients, evidence of intact early visual processing, face sensitivity, fixation, and task engagement across EEG and fMRI suggests that reduced awareness-related markers cannot be attributed solely to failures of attention or sensory input. In contrast, the absence of awareness markers in many severe AD patients was accompanied by a global lack of responsiveness to faces, indicating that null effects in this group may partly reflect impaired sensory processing or task engagement rather than selective loss of awareness.

At the group level, these findings support the notion that perceptual awareness is degraded in AD. However, single-subject analyses revealed substantial variability, including null frontal responses in some healthy controls. This may be related to limited trial numbers, or individual insensitivity to task stimuli during the masking task which constrains the reliability of individual-level inference. Consequently, frontal ignition-like activity or the presence of VAN or LP components cannot yet serve as a reliable biomarker of awareness in individual AD patients.

The implications of the findings of reduced fMRI and EEG markers of awareness for subjective experience remain open. Reduced awareness-related activity in mild-moderate AD may reflect conscious experiences that are weaker, less stable, or less meaningfully integrated with semantic memory, episodic context, and self-referential processing^54–57^. This interpretation is consistent with evidence for impaired feature binding, autonoetic consciousness, and metacognition in AD^6,58,59^Variable degrees of pathology and neurodegeneration within networks, specifically fronto-parietal and default mode networks^47^ may contribute to clinical and phenomenological differences in subjective experience in AD, with heterogeneity in the richness and capacity for higher level awareness, engagement with and reflection on perceptual stimuli. The significantly reduced synchrony during the movie task may reflect differences in the extent to which individuals with AD are able to engage with and experience the movie, with variability in the richness, executive engagement and subjective experience, in contrast to the more uniform experience of healthy control participants. Further studies involving resting state fMRI, DTI and amyloid PET imaging could associate objective markers of awareness and self-reports with quantification of amyloid or tau pathological burden, network integrity and connectivity to assess associations between neurodegeneration and the richness of subjective experience.

Importantly, people with moderate and advanced AD are heterogeneous, with fluctuations in cognition, arousal and apparent awareness both within and between individuals^5,60^. Brief episodes of apparent lucidity in AD patients have been widely reported clinically, underscoring the need for caution in generalising the presence or absence of awareness from single time-point measures or single stimulus modalities^6,61,62^. As awareness is not a unitary phenomenon, the extent of awareness may also vary across sensory domains, according to different ‘objects’ of awareness^63^, with non-visual modalities such as touch or audition potentially playing a more prominent role in later disease stages^64^.

Finally, candidate neural correlates of consciousness must be interpreted in the context of multiple theories of consciousness and ongoing debate about what NCCs are sufficient and necessary for perceptual awareness^51,65,66^. Reduced frontal decoding or absence of ERP components should not be used to categorically infer lack of awareness. GNWT and related frameworks primarily address conscious access and reportability, and diminished ignition may reflect impaired evaluation or reporting rather than absence of experience^67^. Within AD, it may be that progressive neurodegeneration and associated cognitive impairment initially results in a more significant decline in higher-level access consciousness and reflective awareness, with lower-level phenomenological experience akin to sensory registration and experience of the ‘minimal self’^6,68,69^ remaining relatively intact^6,64^. This motivates further research to understand what dimensions and levels of awareness may remain fully or partially intact in advanced dementia, and whether interventions can enhance residual awareness and potentially improve quality of life. Given the clinical heterogeneity and findings of the current study, ethical considerations also demand that reverse inference from apparently null results are avoided. The inherent personhood and value of people with advanced dementia remains paramount, and supportive relationships and social interaction remains important for maintaining selfhood^70^.

In summary, this study provides the first multimodal evidence for reduced markers of perceptual awareness across mild–moderate and severe AD. Reduction of EEG and fMRI neural markers of visual awareness support the view that perceptual awareness becomes progressively compromised in AD. These findings motivate future research to characterise the multiple dimensions, modalities, and temporal dynamics of conscious experience in dementia, with important implications for person-centred care and clinical practice.

## Ethics statement

As participants with severe AD lacked capacity to consent, following the legal framework of the Mental Capacity Act (2005), personal consultees were identified and provided a declaration that the person would have wished to take part in the study (HRA, 2017). The study was approved by the Wales 6 NHS ethics committee (18/WA/0012).

## Acknowledgements

The authors wish to acknowledge the people who gave their time to contribute to this study. The authors gratefully acknowledge the expert support and advice received from the team at the NIHR Join Dementia Research service, which was instrumental to the study. Join Dementia Research is funded by the Department of Health and delivered by the National Institute for Health and Care Research in partnership with Alzheimer Scotland, Alzheimer’s Research UK, and Alzheimer’s Society.

JH is funded by a Wellcome Clinical Research Career Development Fellowship (214547/Z/18/Z). SMF is funded by a Wellcome/Royal Society Sir Henry Dale Fellowship (206648/Z/17/Z) and UKRI under the UK government’s Horizon Europe funding guarantee (selected as ERC Consolidator, grant number 101043666). The Wellcome Centre for Human Neuroimaging is supported by core funding from the Wellcome Trust (206648/Z/17/Z). DB is funded by the Wellcome Trust (210920/Z/18/Z). AMO and SMF are Fellows in the CIFAR Brain, Mind, and Consciousness programme. RH is supported by the NIHR UCLH BRC. LC and JH are supported by the NIHR Applied Research Collaboration South-West.

## Author Contribution Statement

JH, DB, SF, LC, AMO, LN, and RH conceived and planned the experiments. JH and LR carried out the experiments. JH, BB, MM, PAMM, GB processed the experimental data and performed the analysis. All authors contributed to the interpretation of the results. JH took the lead in writing the manuscript. All authors provided critical feedback and helped shape the research, analysis and manuscript.

## Data Availability

Raw data will be deposited in the OpenNeuro registry (openneuro.org), following Brain Imaging Data Structure (BIDS) quality standards. Matlab scripts for analyses of neuroimaging and EEG data will be deposited on the GitHub platform, enabling open access to software code. Requests for access to study data should be made to Dr Jonathan Huntley.

## Supplementary Information

**Supplementary Figure 1:**
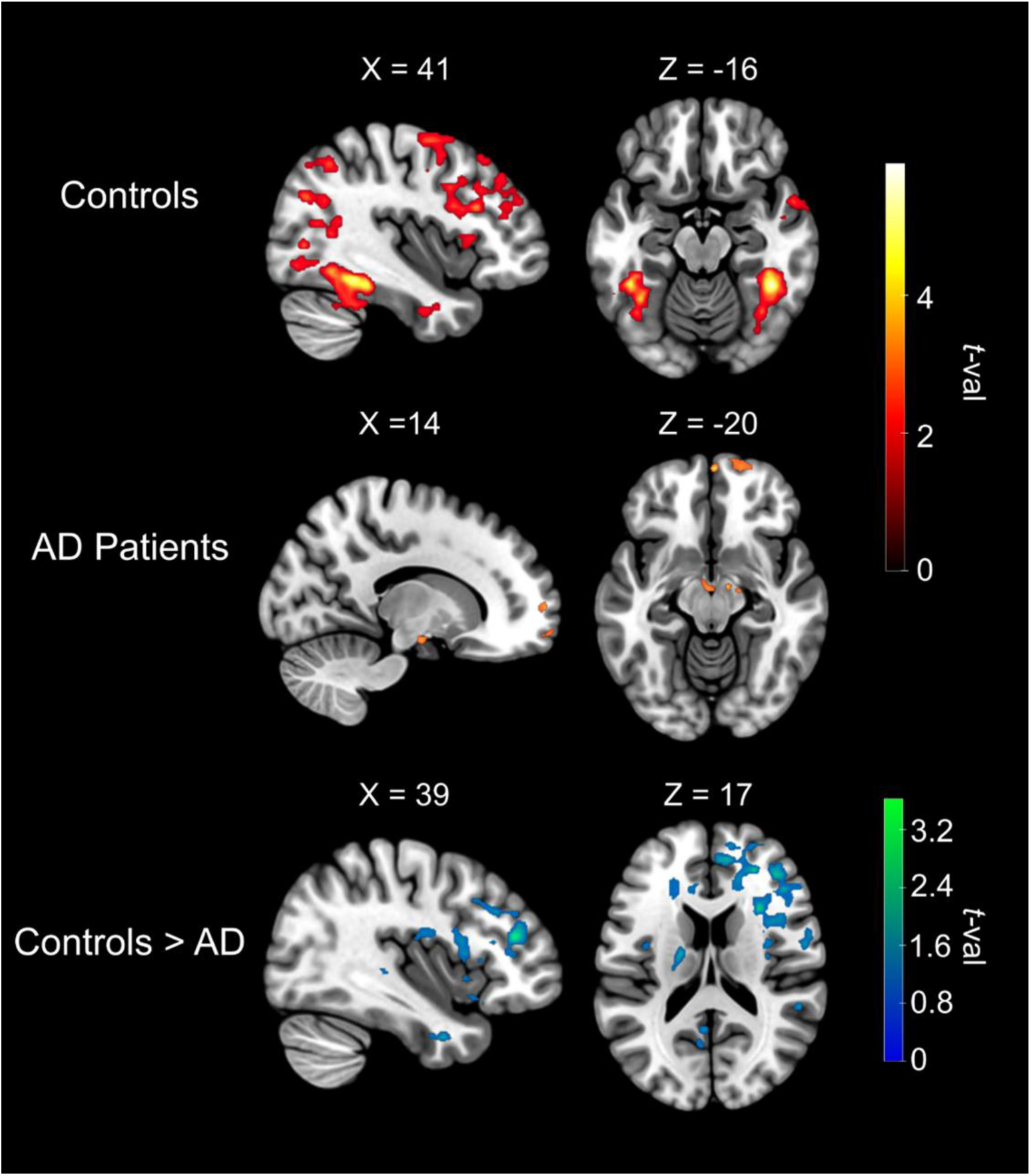
Qualitative visualisation of univariate responses to conscious faces in AD patients and controls. Contrasting trials where participants viewed visible faces vs. subliminal faces reveals neural activity associated with visual awareness. In controls, this contrast was associated with activity in the fusiform gyrus and prefrontal cortex (top panel). Only a limited number of prefrontal voxels were identified as increasing for visible vs. subliminal faces in the AD group, even at the liberal threshold of p <.05 uncorrected (middle panel). Notably, when comparing the activity associated with awareness across the two groups, the controls showed greater frontal activation (bottom panel). Maps are presented at p <.05 uncorrected. Clusters are reported in Supplementary Table 3.

**Supplementary Figure 2:**
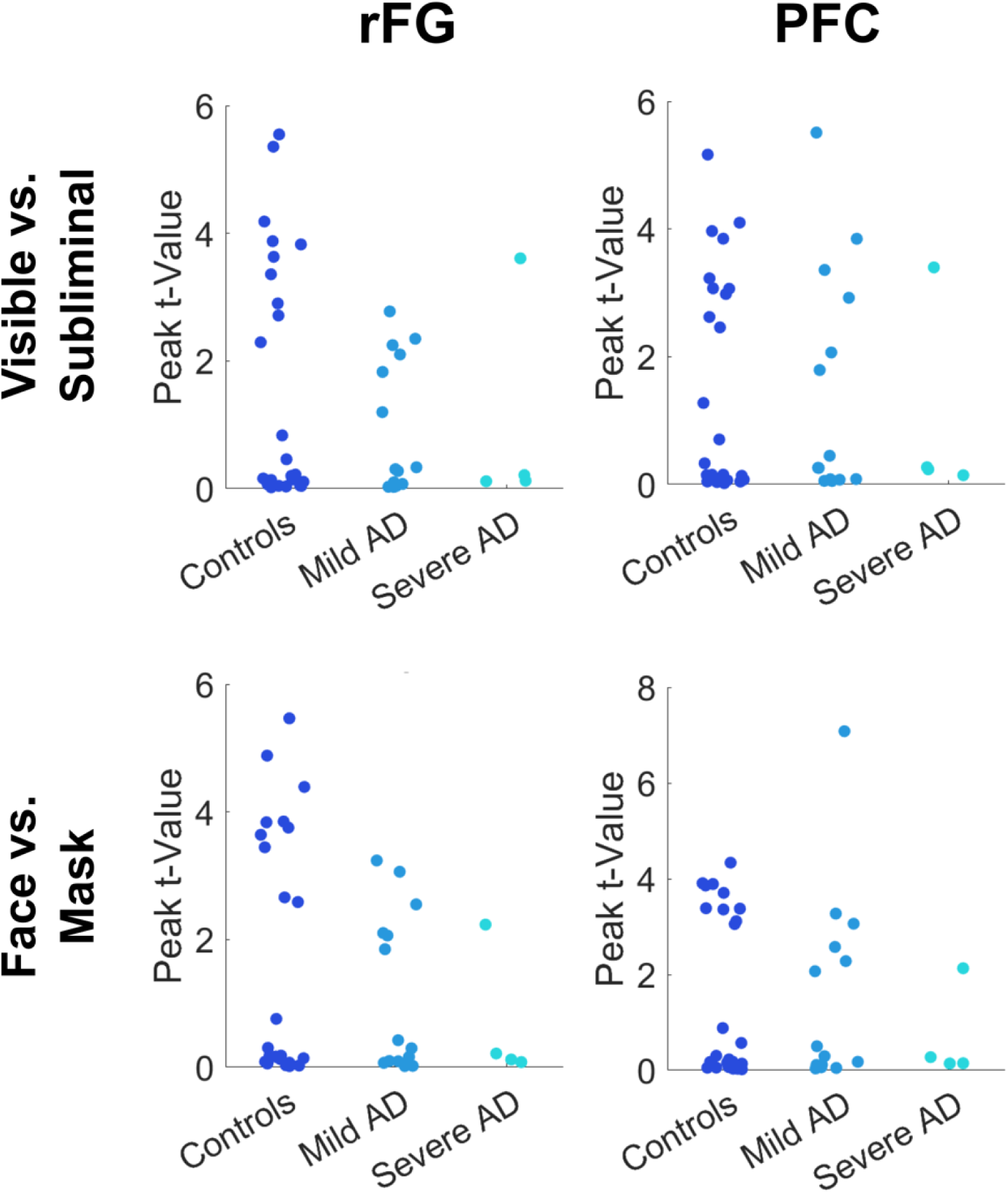
Single subject contrasts of visible vs. subliminal faces in functional ROIs. Top row: Peak t-values from functional ROIs in fusiform and prefrontal regions reveal subjects within each control and patient group who display no increase in activity for visible vs. subliminal faces at the single subject level. As such, a failure to reach statistical significance in such an analysis cannot conclusively be attributed to a patient’s AD diagnosis. Bottom row: Peak t-values from the same ROIs for the visible face vs. scrambled mask contrast. Only one severe AD patient showed sensitivity to faces, suggesting that null findings of activity during may be a result of a global insensitivity to the paradigm, rather than due to patients’ awareness

**Supplementary Table 1.** fMRI Clusters associated with the Visible face vs Mask decoding. XYZ coordinates are MNI coordinates of peak voxel.

| Contrast | Region | Peak Accuracy | No. Voxels | x | y | z |
| --- | --- | --- | --- | --- | --- | --- |
| Controls | Fusiform Gyrus | 0.59 | 4430 | 40 | -50 | -16 |
| Controls | Lateral Occipital Cortex | 0.58 | 2834 | -40 | -78 | 16 |
| Controls | Inferior Frontal Gyrus | 0.55 | 214 | -44 | 8 | 26 |
| Controls | Posterior Cingulate | 0.55 | 202 | -6 | -72 | 16 |
| AD | Supplementary Motor Area | 0.59 | 82 | 8 | -6 | 72 |
| AD | Inferior Parietal | 0.57 | 52 | 52 | -34 | 50 |
| AD | Medial Prefrontal | 0.58 | 42 | -4 | 58 | 14 |
| AD | Lateral Occipital Cortex | 0.57 | 23 | -48 | -60 | -2 |

**Supplementary Table 2:** fMRI clusters where controls show greater Visible Face vs. Scrambled Mask decoding than AD patients. XYZ coordinates are MNI coordinates of peak voxel.

| Region | Peak Difference in Accuracy | No. Voxels | *x* | *y* | *z* |
| --- | --- | --- | --- | --- | --- |
| Intraparietal Sulcus | 0.09 | 92 | -26 | -60 | 54 |
| Posterior Superior Temporal | 0.10 | 75 | 50 | -60 | 10 |
| Lateral Occipital Cortex | 0.09 | 59 | -42 | -86 | 18 |
| Intraparietal Sulcus | 0.09 | 42 | 32 | -64 | 42 |

**Supplementary Table 3.** fMRI Clusters associated with the Visible Face > Subliminal Face group-level contrasts and the subsequent between-group comparison. XYZ coordinates are MNI coordinates of peak voxel.

| Contrast | Region | Peak *t* | No. Voxels | *x* | *y* | *z* |
| --- | --- | --- | --- | --- | --- | --- |
| Controls | Right dlPFC | 4.72 | 2440 | 50 | 26 | 24 |
| Controls | Anterior<br>Cingulate | 4.08 | 973 | 12 | 18 | -18 |
| Controls | Right Fusiform<br>Gyrus | 6.26 | 856 | 42 | -44 | -16 |
| Controls | Left Fusiform<br>Gyrus | 5.02 | 778 | -44 | -42 | -18 |
| Controls | Superior<br>Temporal Sulcus | 4.05 | 630 | 44 | -44 | 14 |
| Controls | Inferior Frontal<br>Gyrus | 4.70 | 521 | -38 | 10 | 28 |
| Controls | Right<br>Intraparietal<br>Sulcus | 3.89 | 411 | 36 | -60 | 42 |
| Controls | Anterior<br>Temporal Lobe | 3.89 | 397 | 46 | 12 | -28 |
| Controls | Left Intraparietal<br>Sulcus | 3.97 | 303 | -36 | -60 | 54 |
| Controls | Corpus Callosum | 3.32 | 261 | -10 | -28 | 26 |
| AD | Right vmPFC | 3.83 | 593 | 2 | 62 | -12 |
| AD | Premotor Cortex | 3.01 | 354 | -16 | 4 | 64 |
| AD | Precentral Sulcus | 3.53 | 326 | 40 | 0 | 36 |
| AD | White Matter | 4.26 | 317 | 36 | -46 | 16 |
| AD | White Matter | 4.02 | 311 | -22 | -42 | 22 |
| AD | Brainstem | 5.94 | 304 | -6 | -14 | -24 |
| AD | Left vmPFC | 3.22 | 275 | -22 | 56 | 6 |
| Controls ><br>AD | Right dlPFC | 3.68 | 1044 | 40 | 38 | 18 |
| Controls><br>AD | Intraparietal<br>Sulcus | 3.84 | 800 | -40 | -40 | 40 |
| Controls><br>AD | Anterior<br>Temporal Lobe | 4.08 | 736 | 62 | 6 | -24 |
| Controls>AD | Parahippocampal<br>Cortex | 3.53 | 486 | 12 | -42 | -10 |
| Controls ><br>AD | Superior<br>Temporal Sulcus | 3.38 | 485 | 52 | -40 | 10 |
| Controls ><br>AD | Cerebellum | 3.35 | 421 | -42 | -52 | -46 |
| Controls ><br>AD | Premotor Cortex | 2.81 | 366 | 52 | -2 | 34 |
| Controls ><br>AD | Brainstem | 3.44 | 350 | 8 | -30 | -16 |
| Controls ><br>AD | Anterior<br>Cingulate | 2.72 | 328 | 10 | 16 | 34 |
| Controls ><br>AD | Inferior<br>Temporal | 3.06 | 320 | -54 | -48 | -18 |
| Controls ><br>AD | Fusiform Gyrus | 3.50 | 318 | -34 | -36 | -24 |
| Controls><br>AD | Putamen | 2.97 | 318 | 24 | -10 | 0 |
| Controls ><br>AD | mPFC | 3.32 | 303 | 26 | 40 | 16 |
| Controls ><br>AD | Cingulate Cortex | 2.84 | 290 | 4 | -14 | 38 |
| Controls ><br>AD | Thalamus | 3.41 | 267 | -14 | -16 | -4 |
| Controls ><br>AD | Posterior<br>Cingulate | 3.14 | 253 | 8 | -36 | 40 |

**Supplementary Table 4.** fMRI Clusters associated with significantly above-chance decoding in searchlight analyses in the Control and AD groups separately. Decoders were trained to classify visible face trials from subliminal face trials and therefore reveal brain regions associated with visual awareness of faces. XYZ coordinates are MNI coordinates of peak voxel.

| **Group** | **Region** | **Peak**<br>**Accuracy** | **No. Voxels** | **x** | **y** | **z** |
| --- | --- | --- | --- | --- | --- | --- |
| Controls | Right<br>Fusiform<br>Face Area | 0.60 | 33,692 | 46 | -54 | -20 |
| Controls | dmPFC | 0.56 | 2337 | 18 | 46 | 36 |
| Controls | dmPFC | 0.56 | 481 | -8 | 54 | 36 |
| Controls | Superior<br>Temporal<br>Sulcus | 0.55 | 329 | 52 | -16 | -10 |
| Controls | vmPFC | 0.55 | 240 | 0 | 26 | -18 |
| Controls | Anterior<br>Insula | 0.55 | 223 | 34 | 20 | -12 |
| Controls | Anterior<br>Temporal<br>Lobe | 0.55 | 208 | 50 | -8 | -32 |
| Controls | dIPFC | 0.55 | 178 | 30 | 10 | 54 |
| Controls | Anterior<br>Temporal<br>Lobe | 0.55 | 177 | -24 | 0 | -30 |
| AD | Posterior<br>Temporal<br>Sulcus | 0.58 | 473 | -58 | -64 | 6 |
| AD | Left<br>Fusiform<br>Gyrus | 0.57 | 172 | -38 | -44 | -18 |
| AD | Occipital<br>Cortex | 0.58 | 152 | -12 | -84 | -20 |
| AD | Inferior<br>Frontal<br>Gyrus | 0.58 | 84 | 56 | 22 | 0 |
| AD | Amygdala | 0.56 | 58 | 16 | 0 | -24 |
| AD | V5/MT | 0.57 | 58 | 56 | -66 | 2 |
| AD | Right<br>Fusiform<br>Face Area | 0.57 | 54 | 40 | -54 | -26 |

**Supplementary Table 5.** fMRI Clusters associated with significantly higher decoding accuracies in the Control group compared to the AD group. XYZ coordinates are MNI coordinates of peak voxel.

| **Region** | **Peak**<br>**Difference**<br>**in**<br>**Accuracy** | **No. Voxels** | ***x*** | ***y*** | ***z*** |
| --- | --- | --- | --- | --- | --- |
| Posterior<br>Temporal Sulcus | 0.10 | 531 | 56 | -66 | 10 |
| Right Fusiform<br>Face Area | 0.12 | 404 | 40 | -48 | -16 |
| Intraparietal<br>Sulcus | 0.09 | 340 | -28 | -66 | 34 |
| Parahippocampal<br>Cortex | 0.09 | 339 | 14 | -66 | 18 |
| Intraparietal<br>Sulcus | 0.09 | 334 | 46 | -54 | 56 |
| Motor Cortex | 0.10 | 326 | 36 | -16 | 70 |
| Intraparietal<br>Sulcus | 0.09 | 266 | 42 | -36 | 46 |
| mPFC | 0.09 | 140 | 24 | 68 | -2 |
| Precuneus | 0.11 | 103 | 6 | -46 | 50 |

**Supplementary Table 6.** fMRI Clusters associated with the Visible Face > Subliminal Face single-subject contrast for patient #1, the only patient with severe AD who showed a statistically significant response to visible faces versus subliminal faces. XYZ coordinates are MNI coordinates of peak voxel.

| Region | Peak *t* | No. Voxels | x | y | z |
| --- | --- | --- | --- | --- | --- |
| Intraparietal<br>Sulcus | 4.11 | 13757 | -26 | -44 | 46 |
| Anterior<br>Insula | 3.51 | 774 | -26 | 32 | 4 |
| Right DLPFC | 3.40 | 1598 | 52 | 18 | 26 |
| Paracingulate<br>Gyrus | 3.28 | 4511 | 14 | 8 | 44 |

**Supplementary Table 7:** Regions with significant correlation between control participants during the movie task. Maps of correlation coefficients for each subject were tested against zero at the group-level in a one-tailed test using SPM12. All group maps were FWE-corrected for multiple comparisons at the cluster level using a cluster forming threshold of 0.001, significant at p < .05

| Volume | Index | Coordinates (x,y,z) | Structure |
| --- | --- | --- | --- |
| 179233 | 13 | 5.4×-59.1×21.2 | 01 |
| 2221 | 9 | 45.9×8.7×29.3 | 11 |
| 1316 | 9 | -48.5×5.8×16.0 | 06 |
| 870 | 11 | 34.1×23.5×6.4 | 05 |
| 774 | 9 | 43.0×36.7×19.7 | 12 |
| 368 | 9 | -36.7×17.6×6.4 | 07 |
| 296 | 9 | 42.2×55.9×-4.7 | 03 |
| 261 | 8 | 51.1×31.6×7.1 | 10 |
| 259 | 8 | 1.7×-26.7×33.0 | 15 |
| 253 | 9 | 44.4×33.0×0.5 | 04 |
| 152 | 8 | -8.7×-14.2×6.4 | 09 |
| 132 | 7 | 54.8×9.4×-17.2 | 02 |
| 129 | 8 | 50.3×22.0×16.7 | 13 |
| 88 | 9 | 10.5×-18.6×6.4 | 08 |
| 60 | 7 | -54.4×1.3×18.2 | 14 |

**Supplementary Table 8:** Regions with significant correlation between mild AD participants during the movie task. Maps of correlation coefficients for each subject were tested against zero at the group-level in a one-tailed test using SPM12. All group maps were FWE-corrected for multiple comparisons at the cluster level using a cluster forming threshold of 0.001, significant at p < .05

| Volume | Index | Coordinates (x,y,z) | Structure |
| --- | --- | --- | --- |
| 2191 | 12 | 0.9×-51.0×44.0 | 8 |
| 292 | 12 | 18.6×-55.5×6.4 | 2 |
| 2487 | 11 | 51.8×-54.0×8.6 | 3 |
| 703 | 10 | -9.4×-70.9×14.5 | 5 |
| 86 | 10 | 1.7×61.1×22.6 | 6 |
| 310 | 9 | 12.0×-70.9×24.1 | 7 |
| 34 | 7 | -30.8×-79.1×10.1 | 4 |
| 73 | 7 | 46.7×-67.3×-2.4 | 1 |

**Supplementary Table 9:** Regions with significant correlation between mild AD participants and the mean activity of the control group during the movie task. Maps of correlation coefficients for each subject were tested against zero at the group-level in a one-tailed test using SPM12. All group maps were FWE-corrected for multiple comparisons at the cluster level using a cluster forming threshold of 0.001, significant at p < .05

| Volume | Index | Coordinates (x,y,z) | Structure |
| --- | --- | --- | --- |
| 43631 | 8 | 11.3×-66.5×11.6 | 03 |
| 5198 | 7 | -47.0×-35.5×41.8 | 10 |
| 271 | 7 | 58.5×-4.6×-19.4 | 01 |
| 91 | 6 | 39.3×-64.3×27.8 | 09 |
| 642 | 6 | 45.9×9.4×30.0 | 08 |
| 305 | 6 | 40.0×-57.7×52.1 | 15 |
| 444 | 6 | 25.3×-39.2×-16.5 | 02 |
| 203 | 6 | 54.8×-2.4×-3.2 | 06 |
| 67 | 6 | -64.7×-48.8×21.9 | 07 |
| 67 | 6 | 59.2×-47.3×-8.3 | 05 |
| 151 | 5 | 49.6×-59.9×44.0 | 13 |
| 182 | 5 | 0.2×-49.6×38.1 | 12 |
| 49 | 5 | -35.2×3.5×29.3 | 11 |
| 164 | 5 | 40.8×-43.7×49.2 | 14 |
| 32 | 5 | 1.7×-57.7×49.9 | 16 |
| 47 | 4 | 29.0×-51.8×-11.3 | 04 |

**Supplementary Table 10:** Regions with significant correlation between the single severe AD participant and the mean activity of the control group during the movie task. Single subject maps of correlation coefficients which were converted to t values (df = (number of timepoints – 2)) and FDR-corrected for multiple comparisons (q = 0.01, p < .05).

| Volume | Index | Coordinates (x,y,z) | Structure |
| --- | --- | --- | --- |
| 6958 | 8 | 0.2×-85.0×16.7 | 5 |
| 4662 | 8 | 61.4×-14.9×-1.0 | 1 |
| 7117 | 7 | -59.5×-23.7×2.0 | 3 |
| 1176 | 7 | -43.3×-69.5×-1.0 | 2 |
| 991 | 6 | 47.4×-64.3×0.5 | 4 |
| 109 | 5 | 43.7×-68.7×16.7 | 7 |
| 344 | 5 | 33.4×-80.5×13.8 | 6 |
| 119 | 5 | 30.4×-64.3×30.7 | 8 |

